# Optimization of magnetic bead-based nucleic acid extraction for SARS-CoV-2 testing using readily available reagents

**DOI:** 10.1101/2021.03.09.21251364

**Authors:** Simon Haile, Aidan M. Nikiforuk, Pawan K. Pandoh, David D. W. Twa, Duane E. Smailus, Jason Nguyen, Stephen Pleasance, Angus Wong, Yongjun Zhao, Diane Eisler, Michelle Moksa, Qi Cao, Marcus Wong, Edmund Su, Martin Krzywinski, Jessica Nelson, Andrew J. Mungall, Frankie Tsang, Leah M. Prentice, Agatha Jassem, Amee R. Manges, Steven J. M. Jones, Robin J. Coope, Natalie Prystajecky, Marco A. Marra, Mel Krajden, Martin Hirst

**Affiliations:** Canada’s Michael Smith Genome Sciences Centre, BC Cancer, Vancouver, British Columbia, Canada; School of Population and Public Health, University of British Columbia, Vancouver, British Columbia, Canada; BC Cancer Research Centre, BC Cancer, Vancouver, British Columbia, Canada; British Columbia Centre for Disease Control Public Health Laboratory, Vancouver, Canada; Department of Microbiology and Immunology, Michael Smith Laboratories, University of British Columbia, Vancouver, British Columbia, Canada; Provincial Laboratory Medicine Services, Provincial Health Services Authority, Vancouver, British Columbia, Canada; Department of Pathology and Laboratory Medicine, University of British Columbia, Vancouver, British Columbia, Canada; Department of Medical Genetics, University of British Columbia, Vancouver, British Columbia, Canada

**Keywords:** SARS-CoV-2, COVID-19, nucleic acid extraction, RNA, cross-well, contamination, Hamilton NIMBUS, Magnetic beads

## Abstract

The COVID-19 pandemic has highlighted the need for generic reagents and flexible systems in diagnostic testing. Magnetic bead-based nucleic acid extraction protocols using 96-well plates on open liquid handlers are readily amenable to meet this need. Here, one such approach is rigorously optimized to minimize cross-well contamination while maintaining sensitivity.

**Article Summary:** A scalable, non-proprietary, magnetic bead-based automated nucleic acid extraction protocol optimised for minimum cross-well contamination

---

The COVID-19 pandemic has placed unprecedented strain on instrument and consumable supply chains for SARS-CoV-2 nucleic acid (NA) testing (1). NA protocols involve lysis and purification of NAs on columns or magnetic beads. Bead-based protocols are amenable to automated workflows and are widely available, rendering them attractive alternatives to proprietary commercial offerings (2). Recent reports have suggested that generic bead-based protocols can be successfully deployed on generic open-deck liquid handling instruments (3-5); yet lack rigorous measures of specificity and sensitivity that are required for clinical deployment.

## The Study

We sought to establish an automated protocol to support extraction-based SARS-CoV-2 NA testing using generic reagents, automated on an open deck Hamilton NIMBUS96 liquid handler. We benchmarked this against an existing clinical NA testing workflow in place at the BC Centre for Disease Control Public Health Laboratory (BCCDC PHL) (Vancouver, Canada) that relies on the MagMax™ −96 Viral RNA Isolation kit (Thermo Fisher Scientific) deployed on the Applied BioSystems MagMax Express™ 96 platform, hereafter referred to as the “MagMax”. MagMax features bead-bound NAs that are transferred serially to five plates containing wash solutions and elution buffer via 96 magnetized rods with disposable sheathes. Widespread adoption of this commercial workflow during the pandemic has driven ongoing shortages in reagents and motivated this study. This reagent shortage extended to many other manufacturers of automated NA extraction systems.

Magnetic bead-based NA purification workflows deployed on open liquid handling platforms support various chemistries and plastic-ware configurations, providing critical flexibility in the face of global supply chain instabilities. Here, we provide a benchmarked standard operating procedure (SOP; **Appendix 1 and 2**) that employs a guanidine-thiocyanate containing lysis buffer followed by NA purification using magnetic beads and deployed on a Hamilton NIMBUS liquid handler (**Appendix-3A**). The SOP is designed to accept specimens aliquoted into a plate from a variety of commercial transport mediums (Copan UTM, Hologic STM, Roche cobas® PCR Media, YOCON UTM) or common laboratory buffers.

Cultured Influenza A virus (Flu-A) spiked into transport medium was used for initial comparisons between the NIMBUS and MagMax protocols. Following extraction, Flu-A RNA recovery was measured using a TaqMan qRT-PCR assay, developed by BCCDC, that detects Flu-A, Flu-B and RSV. The PCR cycle threshold (Ct) values obtained from the NIMBUS protocol were lower than those obtained from the MagMax protocol across dilutions when Copan UTM was used as virus diluent by an average of 1.13 Ct (*p*=0.0039; **Appendix 3B**). When Hologic STM was used, the opposite was observed, with higher values from the NIMBUS protocol compared to those from the MagMax protocol by an average of 0.22 Ct (*p*=0.0301; **Appendix 3C**). These results suggest that the NIMBUS protocol provides comparable sensitivity to the MagMax protocol.

To measure specificity of our protocol we deployed a “checkerboard” input plate where Copan UTM containing Flu-A was alternated with Copan UTM alone. This test revealed a ∼85% specificity; carryover into blank wells was judged to be an aggregate effect of the manual processes performed in the biosafety cabinet (BSC) and automated liquid handling. To decouple these sources of contamination, we utilized synthetic DNA (g-block) controls and matched primers and probe sets. A master checkerboard plate was first generated by aliquoting an amount of g-block DNA sufficient for an extraction-free control plate and three extraction test plates. Using a dedicated NIMBUS, the master plate was aliquoted into a control plate containing elution buffer only and into a deep-well plate that was pre-loaded with a mixture of Copan UTM, RLT Plus, beads, and isopropanol to mimic the extraction chemical milieu (extraction plates), respectively. The samples from two of the extraction plates were then purified using a second NIMBUS and those from the third extraction plate were purified on a third NIMBUS. As shown in **Figure 1**, all the 40 blank wells in the control plate were determined to be negative via the qPCR assay (i.e. undetermined Ct values) (100% specificity). In contrast, the three extraction plates displayed 17, 13 and 12 false positive wells, representing 65% specificity.

**Figure 1.**
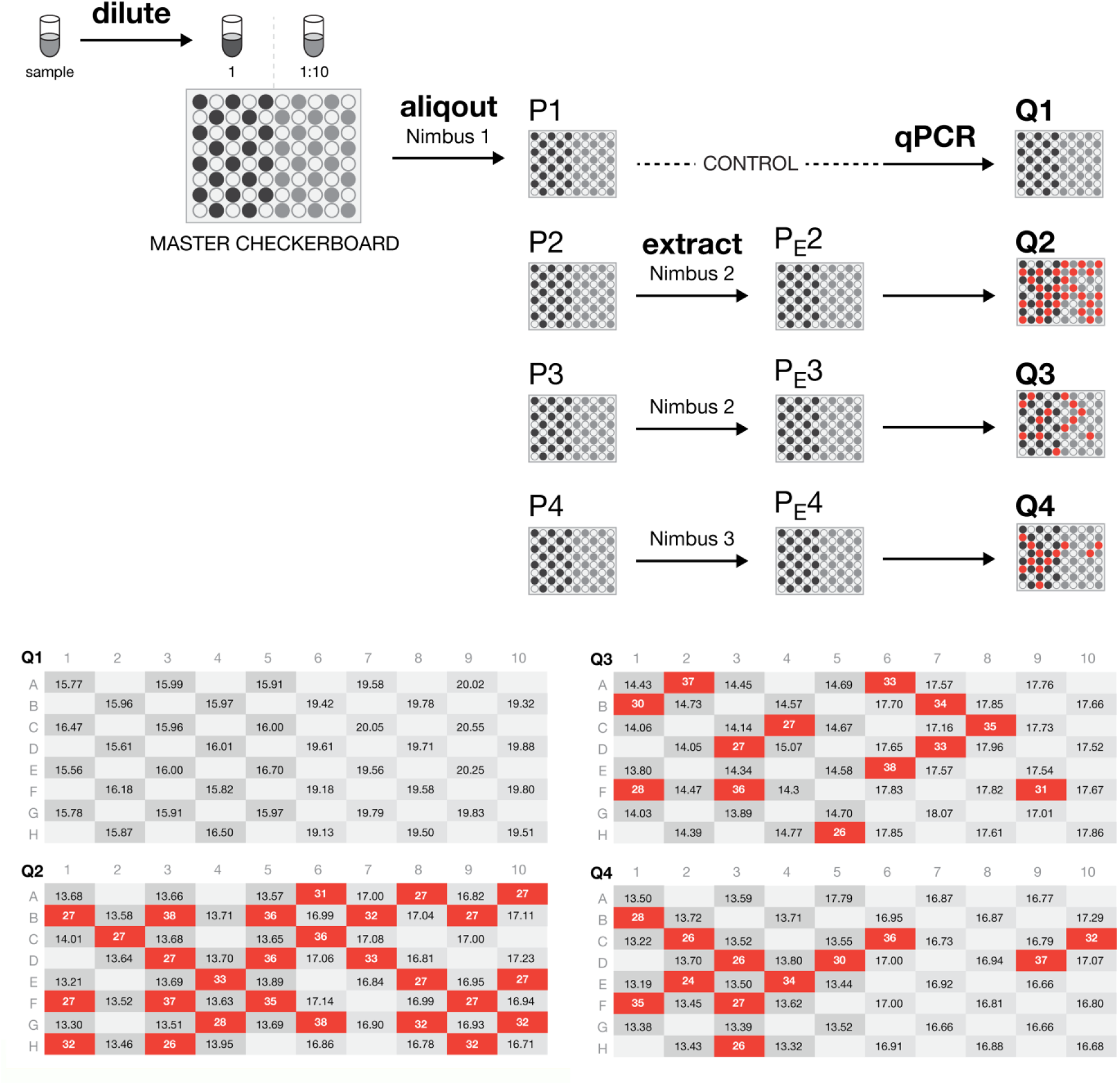
Assessment of cross-well contamination associated with a liquid handler. The contamination assessment Workflow is shown in the upper panel. This assay is designed to decouple the manual upstream BSC steps from the steps on the liquid handler (in this case a NIMBUS). A synthetic DNA fragment (g-block) is used as a starting material and a master g-block checkerboard plate is manually generated. The g-block DNA is aliquoted into a plate that contains elution buffer (control plate) and a deep-well plate that was pre-loaded with a mixture of Copan UTM, RLT Plus, beads, and isopropanol (extraction plates), respectively, using a devoted liquid handler. The samples from the extraction plates are then purified on separate liquid handlers. The g-block DNA eluates from the extraction plates and diluted g-block DNA from the control plates are subsequently used as templates in the same run of qPCR (lower panel).

To improve specificity, we switched from 1.2mL to 2.2 mL deep-well plates, reduced tip mixing steps and number of washes and optimized pipetting techniques to eliminate residual droplets. We wrote new code to eliminate extraneous vertical movement of the robot head between aspirate and dispense steps and to reduce the robot gantry’s lateral speed to prevent dislodging of any residual droplets adhering to tips. The details of all the changes are described in **Appendices 4 and 5**.

We tested the aggregate effects of all the changes in the optimized NIMBUS protocol (**Figure 2A**) against the MagMax protocol using a Flu-A dilution series in Copan UTM. The optimized NIMBUS protocol demonstrated increased sensitivity compared to MagMax by an average of 0.98 Ct (*p*=0.000015; **Figure 2B)**.

**Figure 2.**
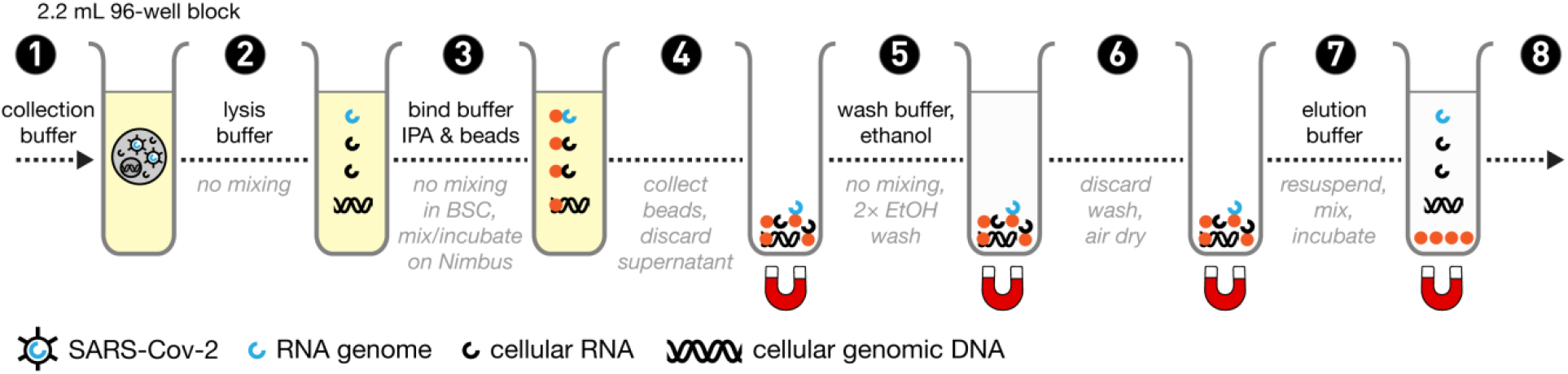
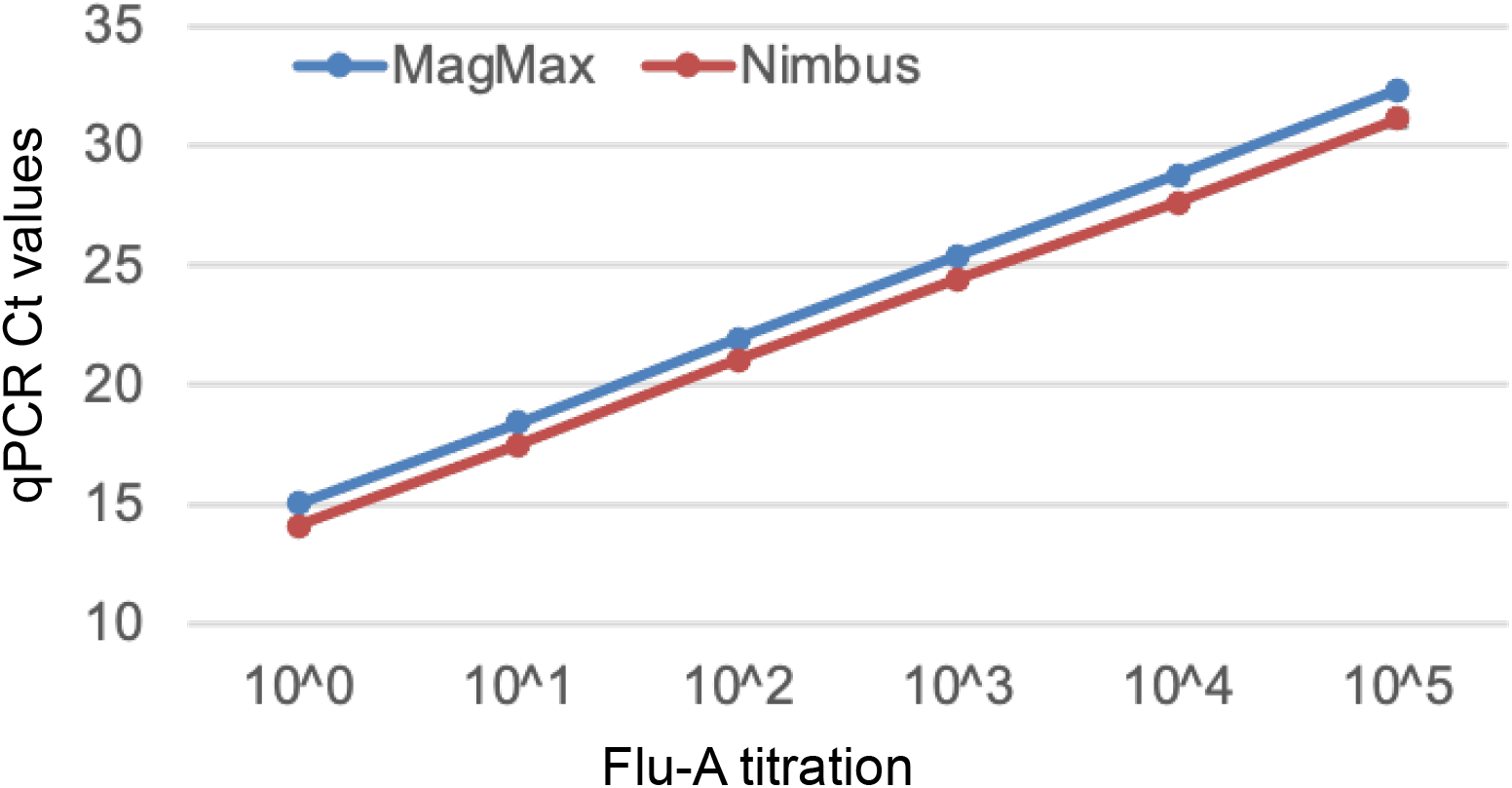
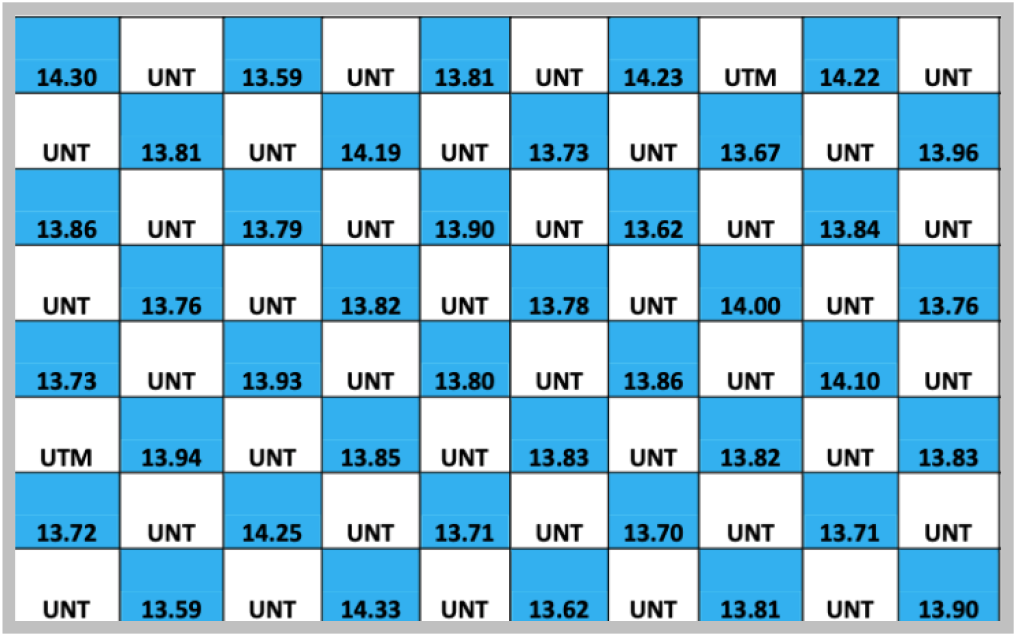
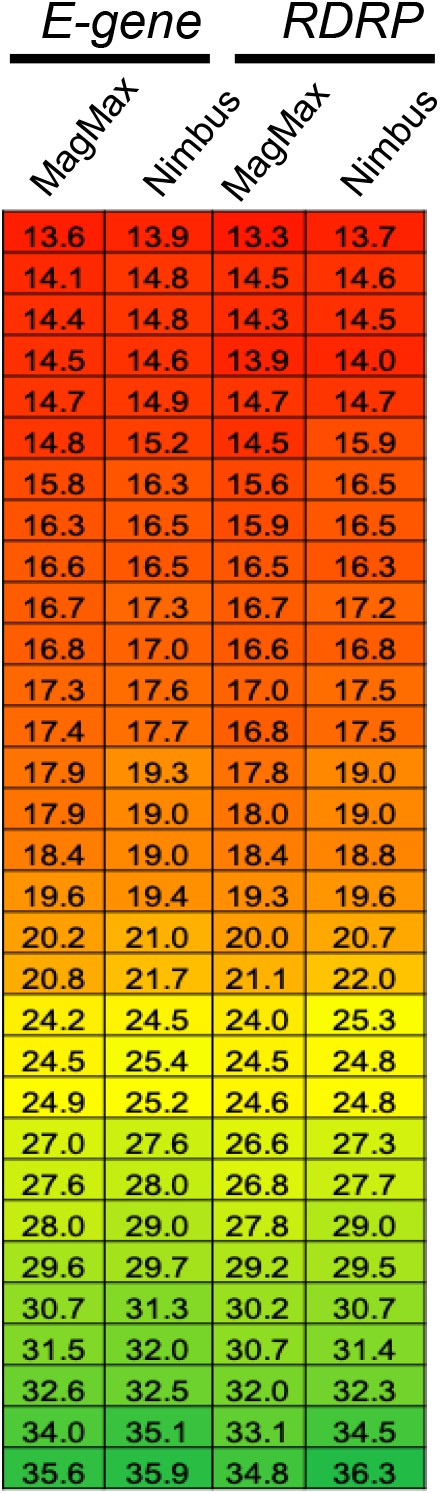
Specificity and sensitivity of the optimized NIMBUS-based protocol. (**A**) Workflow of the optimized NIMBUS-based protocol. Modifications included: removing the manual mixing steps in the BSC (2), performing all NIMBUS steps in 2.2 mL plates instead of 1.2 mL (1-8); removing the mixing step following addition of the wash buffer on NIMBUS (5), reducing the number of ethanol washes (5), and reducing the number of mixing in the elution step (7). The other changes that were implemented are summarized in Appendix 6-7. (**B**) Comparison of sensitivity of the NIMBUS protocol with that of the MagMax. One to five log dilutions of Flu-A virus stocks were spiked into Copan UTM and yield was measured via qRT-PCR. *p*=0.000015 (paired, two-tailed t-test; n=4 for each of the dilutions except 10^5 NIMBUS; n=3) error bars=standard deviations. (**C**) Assessment of cross-well contamination levels associated with the NIMBUS protocol. Checkerboard pattern was set-up with alternating wells of Flu-A virus that was spiked into Copan UTM and Copan UTM without Flu-A virus. UNT=undetected. (**D**) Comparison of the improved NIMBUS-based protocol with the MagMax protocol using Covid-19 samples. A heatmap on Ct values obtained from qRT-PCR measurements for both the *RDRP* and *E-gene* targets is shown. These data are from two independent experiments.

Having established comparable sensitivity of our optimized NIMBUS protocol to that of the initial version of the NIMBUS and the MagMax protocols, we next tested its specificity. We performed three independent experiments using g-block checkerboards, each including two extraction-free control plates and two extraction test plates (total of 240 positive wells and negative wells). None of the blank wells had detectable NA and all wells with g-block DNA yielded expected Ct values indicating 100% sensitivity and specificity. A Flu-A checkerboard was performed and again achieved 100% sensitivity and specificity (**Figure 2C**).

Final benchmarking with nasopharyngeal swabs samples for SARS-CoV-2 testing was performed to compare the NIMBUS to the MagMax protocol. Two independent experiments were run with 34 negative and 31 positive SARS-CoV-2 samples. Results show concordance between the protocols, each detecting 34 negatives and 31 positives. For the positive samples with Ct values that ranged from 13.8 to 36.8 (**Figure 2D**), there was a strong correlation between the two protocols (R^2^>0.996) (**Appendix-6**). There were slightly higher Ct values (by a median of 0.43 Ct for *RDRP* and 0.50 Ct for *E-gene*) from the NIMBUS protocol (*p=*1.89E-08 and *p*=9.94-E08, respectively). Taken together, our analysis indicates that the optimized NIMBUS protocol yields 100% specificity and sensitivity and comparable RNA yield, compared to the MagMax protocol used routinely at BCCDC PHL.

## Conclusion

Generic nucleic-acid purification protocols provide an alternative reagent stream for SARS-CoV-2 testing but require optimization and customization to meet clinical sensitivity and specificity requirements. Here we provide a benchmarked SOP for one such protocol deployed on a Hamilton NIMBUS platform.

## Supporting information

Appendix 1 and 2 - SOPs

## Data Availability

Data may be requested

## Acknowledgments

Support for this work was provided by contributions from the Provincial Health Services Authority, Genome BC/Genome Canada (202SEQ, 212SEQ, 12002, COV-088), the Canada Foundation for Innovation (20070, 30981, 30198) and the BC Knowledge Development Fund.

## Appendix Figure legends

**Appendix 3. First iteration of the NIMBUS-based protocol**. (**A**) Workflow of the Iteration-1 of the NIMBUS-based protocol. The upstream part of the process (blue boxes) is performed manually within the biosafety cabinet (BSC). The downstream steps are performed on the NIMBUS (grey boxes). (**B**) Comparisons with the MagMax protocol. Dilutions of Flu-A virus stocks were spiked into the sample collection, the Aptima Specimen Transfer Medium manufactured by Hologic (STM), followed extraction using the MagMax or NIMBUS protocols. *p*=0.0039 (paired, two-tailed t-test; excluding the data points for the most diluted sample); n=1 for each dilution point; (**C**) The same as in (B) but with Copan Universal Transport Medium (UTM) instead of Hologic STM. *p*=0.031 (paired, two-tailed t-test); n=3 for each dilution point; error bars=standard deviations.

**Appendix 4**. Workflow of the improved version of the NIMBUS-based protocol. Modifications are indicated in red text and included: removing the manual mixing steps in the BSC, performing all NIMBUS steps in 2.2 mL plates instead of 1.2 mL; removing the mixing step following addition of the wash buffer on NIMBUS, reducing the number of ethanol washes, and reducing the number of mixing in the elution step. The other changes that were implemented are summarized in Appendix 5.

**Appendix 5**. Improvements of the NIMBUS-based protocol to reduce cross-well contamination levels. Modifications of the liquid handling techniques and other aspects of automation are listed. The other changes that were implemented are summarized in Appendix 4.

**Appendix 6**. Comparison of the improved NIMBUS-based protocol with the MagMax protocol using Covid-19 samples. A linear regression on Ct values obtained from qRT-PCR measurements for both the *RDRP* and *E-gene* targets is shown. Insert includes correlation values (R^2^). These data are from two independent experiments. *p=*1.89E-08 and *p*=9.94-E08 for *RDRP* and *E-gene*, respectively (paired, two-tailed t-test).

