## Appendix 1 and 2 - SOPs for "Optimization of magnetic bead-based nucleic acid extraction for SARS-CoV-2 testing using readily available reagents"

**Standard Operating Procedure - Total nucleic acid extraction from nasopharyngeal swab samples**

### I. Purpose

To enable extraction of total nucleic acids from nasopharyngeal swab samples for testing and other applications related to SARS-CoV-2 and Influenza A/B.

### II. References

| Document Title | Document Number |
|---|---|
| Agencourt RNAdvance Tissue Protocol (Note: ALINE EvoPure RNA Isolation Kit uses the same protocol as Agencourt RNAdvance Tissue Protocol). | 000473v003 |

### III. Safety

All Laboratory Safety procedures will be complied with during this procedure. The required personal protective equipment includes a laboratory coat and gloves. For chemical safety, see safety data sheet (SDS).

### IV. Materials and Equipment

| Name | Supplier | Number |
|---|---|---|
| 2.2 mL KingFisher Deep Well 96 Plate | Thermo Scientific | 95040450 |
| EvoPure RNA Isolation Kit | ALINE | R-907-400-C5 |
| RLT Plus | Qiagen | 1053393 |
| IPA (2-Propanol) 4 L | Fisher | A464-4 |
| MicroAmp Clear Adhesive Film | ABI | 4306311 |
| Disposable Trough | VWR | 89094-664 |
| Multi-channel pippette | | |
| Tips for multichannel | | |
| Single channel Pipette | | |
| Tips for single channel | | |
| 50 mL Falcon tubes | | |
| Bleach | | |
| 1000 µL Rainin tips | Rainin | RT-L1000F |
| 2.2 mL KingFisher 96 Deep Well Plate | Thermo Scientific | 95040450 |
| EvoPure RNA Isolation Kit | ALINE | R-907-400-C5 |
| Anhydrous Ethyl Alcohol (100% Ethanol) | Commercial Alcohol | 00023878 |
| AB1000 96-well 200 µL PCR plate | Fisher | AB1000 |
| Bench Coat (Bench Protection Paper) | Fisher | 12-007-186 |
| Black ink permanent marker pen | VWR | 52877-310 |
| Ultrapure Water, Dnase/Rnase-free, distilled | Invitrogen | 10977-023 |
| Disposable Trough | VWR | 89094-664 |
| Safetouch gloves | Fisher | 270-058-53 |
| Gilson P1000 pipetman | Mandel | GF-23602 |
| Gilson P200 pipetman | Mandel | GF-23601 |
| Multi-channel Pipette at BCCDC | | |
| Pipette Tip, Filter, Sterile | | |
| Hamilton Microlab NIMBUS96 | Hamilton | |

| | | |
|---|---|---|
| Ice bucket – Green | Fisher | 11-676-36 |
| IPA (2-Propanol) 4 L | Fisher | A464-4 |
| Large Kimwipes | Fisher | 06-666-117 |
| Large Volume Magnet Plate | Alpaqua | 96M-EX |
| Mandel P1000 DF1000 tips | Mandel | GF-F171703 |
| Mandel P200 DF200 tips | Mandel | GF-F171503 |
| MicroAmp Clear Adhesive Film | ABI | 4306311 |
| NIMBUS 1000 µL CO-RE filter tips | Hamilton | 235821 |
| NIMBUS 300 µL CO-RE filter tips | Hamilton | 235832 |
| Peltier Heaters | In-house | N/A |
| Reservoir, 96 well pyramid, 287 mL | Agilent | 201244-100 |
| RNAse free 1.5 mL Eppendorf tube | Ambion | 12400 |
| RNAse Zap | Ambion | 9780 |
| Small Autoclave waste bags 10”X15” | Fisher | 01-826-4 |
| VX-100 Vortex Mixer | Rose Scientific | S-0100 |
| Wet ice | In house | N/A |
| Reservoir, Pyramid PP 86 mL | Agilent | 201254-100 |
| Autoclave waste bags 12" x 24" | ThermoFisher | 01-826-5 |
| Heavy Foil Tape Roll | Scotch/3M | 34000740 |

### V. Procedure: Extraction of Total Nucleic Acids

#### 1. Reagent Preparation Guidelines

1.1. RNases are ubiquitous and general precautions should be followed to avoid the introduction of contaminating nucleases.

1.2. Always work with gloved hands.

1.3. Use RNase-free, filtered pipette tips for pipetting.

1.4. Use reagents, plastic lab ware, and other disposable consumables that are certified to be RNase-free.

1.5. Use dedicated RNase-free equipment, e.g. pipettes, etc.

1.6. When available, work in a separate space designated for RNA work.

1.7. When appropriate, prepare small individual aliquots of reagents and consumables to avoid repeated transfer out of stock buffers. This lowers the risk of contaminating the stock solution.

1.8. Wipe down work surfaces with RNAse Zap before starting.

1.9. Use appropriate positive and negative controls.

### 2. Workflow Overview

The workflow of the extraction process is depicted below. The upstream part of the process (in blue; steps 1-3) is performed within a class II Biological Safety Cabinet (BSC). The downstream steps are performed on the Microlab NIMBUS (Hamilton) liquid handling platform (in grey; steps 4-7). The upstream process begins with transferring a portion of the sample from the Copan UTM or Hologic STM tubes into a 2.2 mL KingFisher Deep 96-well plate. Qiagen's RLT Plus lysis buffer is then added to the samples. This step serves to lyse the sample and contributes to viral inactivation. Subsequently, a mixture of magnetic beads in isopropanol is added to the lysate to capture of nucleic acid from the lysate on the magnetic beads for subsequent purification and the isopropanol facilitates the second part of inactivating residual (non-lysed) virus. Following these steps within the BSC, the plate with inactivated virus is taken to the NIMBUS deck outside the BSC as outlined.

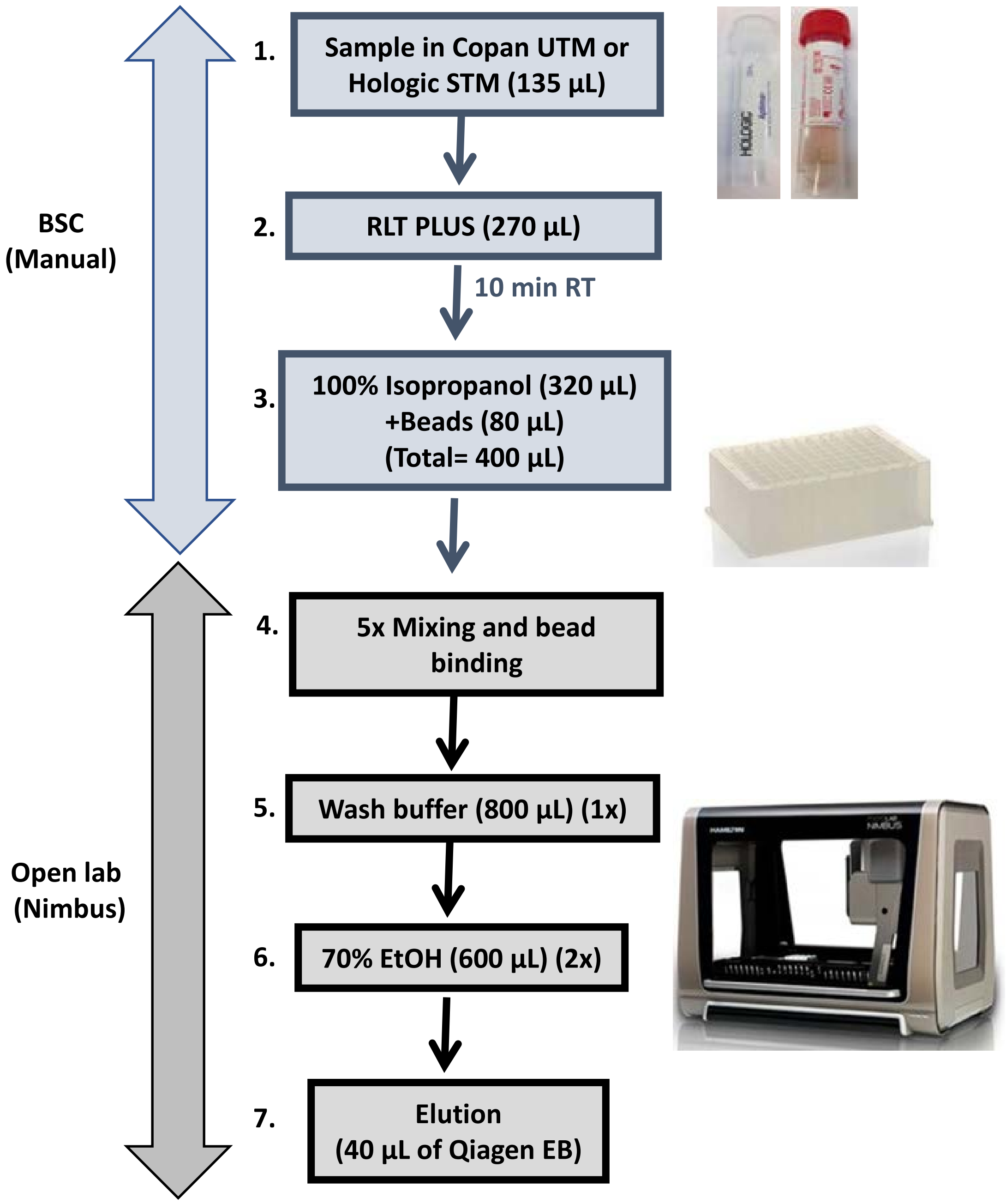

BSC
(Manual)
1.
Sample in Copan UTM or Hologic STM (135 µL)
2.
RLT PLUS (270 µL)
10 min RT
3.
100% Isopropanol (320 µL)
+Beads (80 µL)
(Total= 400 µL)
Open lab
(Nimbus)
4.
5x Mixing and bead binding
5.
Wash buffer (800 µL) (1x)
6.
70% EtOH (600 µL) (2x)
7.
Elution
(40 µL of Qiagen EB)

### 3. Sample transfer, lysis, and addition of bead binding mix

3.1. Before beginning the extraction protocol:

3.1.1. In a clean area, prepare bind mix as follows using one 50 mL Falcon tube per plate:

| **Solution** | **1 rxn (no dead volume)** | **1 plate (with dead volume)** | **2 plates (with dead volume)** |
|---|---|---|---|
| Tissue RNA bind beads | 80 µL | 9.6 mL | 9.6 mL *2 |
| Isopropanol | 320 µL | 38.4 mL | 38.4 mL*2 |
| **Total volume** | **400** µL | **48 mL** | **48 mL*2** |

3.2. Bring RLT Plus, the bind mix tubes and required number of 2.2 mL KingFisher Deep Well 96 Plate to the BSC and set up the following steps in the BSC.

3.3. In a BSC, remove lid of racked samples tubes one at a time.

3.4. Transfer **135 µL of sample** to the 2.2 mL KingFisher Deep Well 96 Plate keeping track of sample-well map. Dip the tip into bleach to kill virus before disposing of tip in a biosafety container.

3.5. Pour RLT Plus into a reagent trough.

3.6. Carefully, without splashing, transfer **270 µL RLT Plus** to each well using a multichannel pipette one column at a time. Do not mix. Change tips between columns.

3.7. Double check the plate to ensure RLT plus is added to all wells and ensure the top edge of the wells is dry and clean. Seal the plate with clear tape.

3.8. Incubate for 10 minutes at room temperature.

3.9. Shake the prepared **bead bind mix** in the Falcon tube(s), pour into a reagent trough, and immediately transfer **400 µL** to each well using a multichannel pipette one column at a time carefully without causing any splashing. Do not mix. Change tips between columns.

3.10. Double check the plate to ensure that the bead mix is added to all wells and ensure the top edge of the wells is dry and clean. Seal the plate with a clear tape. Wipe the plate with bleach, incubate 10 minutes inside BSC, and wipe with 70% ethanol.

3.11. Carefully transport the plate(s) to the NIMBUS workstation.

### 4. Purification of total nucleic acid

4.1. Prepare a working Wash Buffer by adding Isopropanol to one bottle of Wash Buffer concentrate (Part of the EvoPure extraction kit). Follow the instructions on the bottle for the specific amount of Isopropanol that is required.

4.2. Pour 70% ethanol and working wash buffer, respectively, into respective high profile reservoir plates (Reservoir 96 well pyramid pp 287 mL) and cover them with a clear tape seal until use.

| **Solution** | **1 well (no dead volume)** | **1 plate (with dead volume)** | **2 plates (with dead volume)** |
|---|---|---|---|
| 70% ethanol for plate 1 | 600 µL*2X | 170 mL | NA |
| 70% ethanol for plate 2 | 600 µL*2X | 170 mL | NA |
| Working Wash Buffer for 1 or 2 plates | 800 µL*1X | 110 mL | 220 mL |

4.3. Pour 30 mL of Qiagen's elution buffer into a low profile reservoir plate (Plate, 384-Well reservoirs, diamond-bottom, Low-Profile) and cover it with clear tape seal until use.

4.4. Retrieve the 2.2 mL KingFisher Deep Well 96 Plate containing 805 µL of lysate and bead binding mix from step 3.11.

4.5. Log onto the NIMBUS and open the software (Hamilton Run Control):

> <u>NIMBUS</u>: C: > Program Files (x86) > Hamilton Company > Methods > Production > Covid-19 2-Plate Extraction v4.1

4.6. The NIMBUS allows the processing of either 1 or 2 plates at time; you will be prompted to choose.

4.7. Follow prompts on the NIMBUS for the rest of the procedure. Note that there will be two layers of the deck layout that the NIMBUS will specify.

4.8. Note that this is not a walk-away program; users will need to attend to tip needs, movement of plates to save tips, and other aspects as prompted by the NIMBUS.

4.9. Following elution of the total nucleic acids, seal the destination plate and perform a quick spin. For temporary storage, cover with clear tape seal and place the plate on ice. For longer term storage, use heavy foil tape and store at -80°C.

4.10. Clear the deck of all plastic labware and discard. Discard any unused reagents. Log off from the computer.

**Manual total extraction of total nucleic acid from nasopharyngeal swab specimen**

*Refer to workflow, materials and guidelines above*

1. Before beginning the extraction protocol:

   1.1. Retrieve sufficient 70% Ethanol (EtOH) from 4°C and to warm up to room temperature. You will need 1.9 mL/sample.

   1.2. Retrieve the 2.2 mL KingFisher Deep Well 96 Plate containing 805 µL of lysate and bead binding mix

2. Using a multi-channel pipettor Mix 5 times at 96% of the total volume of lysate and bead binding mix (775 µL).

3. Incubate at room temperature for 5 minutes.

4. Place the sample plate onto a Magnum FLX magnet plate for 10 minutes minimum or longer if needed until the supernatant has cleared.

5. Remove and discard supernatant.

6. While the sample plate is on the magnet, add 800 µL of Wash I Buffer to each sample.

7. Allow the beads to settle for 2 minutes.

8. Remove the wash buffer.

9. Dispense 70% EtOH into a fresh V-bottomed sterile solution basin. With the plate on the magnet, add 600 µL of 70% EtOH to each sample. Allow beads to resettle for 2 minutes. Remove all EtOH.

10. Repeat the EtOH wash, for a total of 2 EtOH washes.

11. With the plate on magnet, allow to dry for 10 minutes. During the incubation, dispense Qiangen’s EB buffer into a fresh fresh V-bottomed sterile solution basin.

12. Remove the plate from the magnet and add 40 µL of EB buffer.

13. Place a spacer on the magnet

14. Move the plate to the magnet with the spacer on and center the beads for 1 minute.

15. Move the plate off the magnet.

16. Mix 10 times or until beads are fully resuspended.

17. Allow elution incubation for 7 minutes.

18. Remove the spacer from the magnet.

19. Move the plate back onto the magnet without the spacer on for 2 minutes to clear the supernatant.

20. Transfer the supernatant (containing the total nucleic acids) to an AB1000 96-well plate, the final destination plate.

21. Seal the plate and perform a quick spin. For temporary storage, seal with a clear tape seal and place the plate on ice. For longer term storage, use foil tape and store at -80°C.

### APPENDIX 2

#### Detailed summary of robot steps

Customized Nimbus Code as well as .STEP or .SLT files of the plate spacer described below are available upon request

1. Mix 5x at 775 µL.
2. Incubate at room temp for 5 minutes.
3. Place on Magnum FLX magnet plate for 10 minutes.
4. Pipette off and discard all 800 µL supernatant (over-aspirate volume 820 µL is 20 µL more than the volume in the wells).
5. With the plate on the magnet, add 800 µL of prepared wash buffer from step 3.1.
6. Allow beads to resettle for 2 minutes.
7. Remove all wash buffer with an over-aspirate of 803 µL.
8. With the plate on the magnet, add 600 µL of 70% EtOH. Allow beads to resettle for 2 minutes. Remove all EtOH with an over-aspirate of 613 µL.
9. Repeat this EtOH wash, for a total of 2 EtOH washes.
10. Move the plate off the magnet.
11. Allow to dry for 10 minutes.
12. Pour EB buffer into a low profile reservoir and place on deck as prompted. The robot will prompt you to remove the waste plate and place this reservoir on the deck.
13. The robot will prompt you to swap the P1000 tips for P300 tips
14. Add 40 µL of EB buffer.
15. The robot will prompt you to place a spacer on the magnet.
16. Move the plate to the magnet with *spacer on for 1 minute to center the beads.
17. Move the plate off the magnet.
18. Mix 25 times at 25 µL.
19. The robot will prompt you to remove the spacer.
20. Move the plate to the magnet without spacer and clear for 2 minutes.
21. Robot will prompt to put destination plate on the deck.
22. Transfer the 40 µL supernatant (containing the total nucleic acids) to an AB1000 96-well plate, the final destination plate, using an over-aspirate volume of 50 µL.
23. Seal the plate and quick spin. For temporary storage, seal with a clear tape seal and place the plate on ice. For longer term storage, use heavy foil tape and store at -80°C.

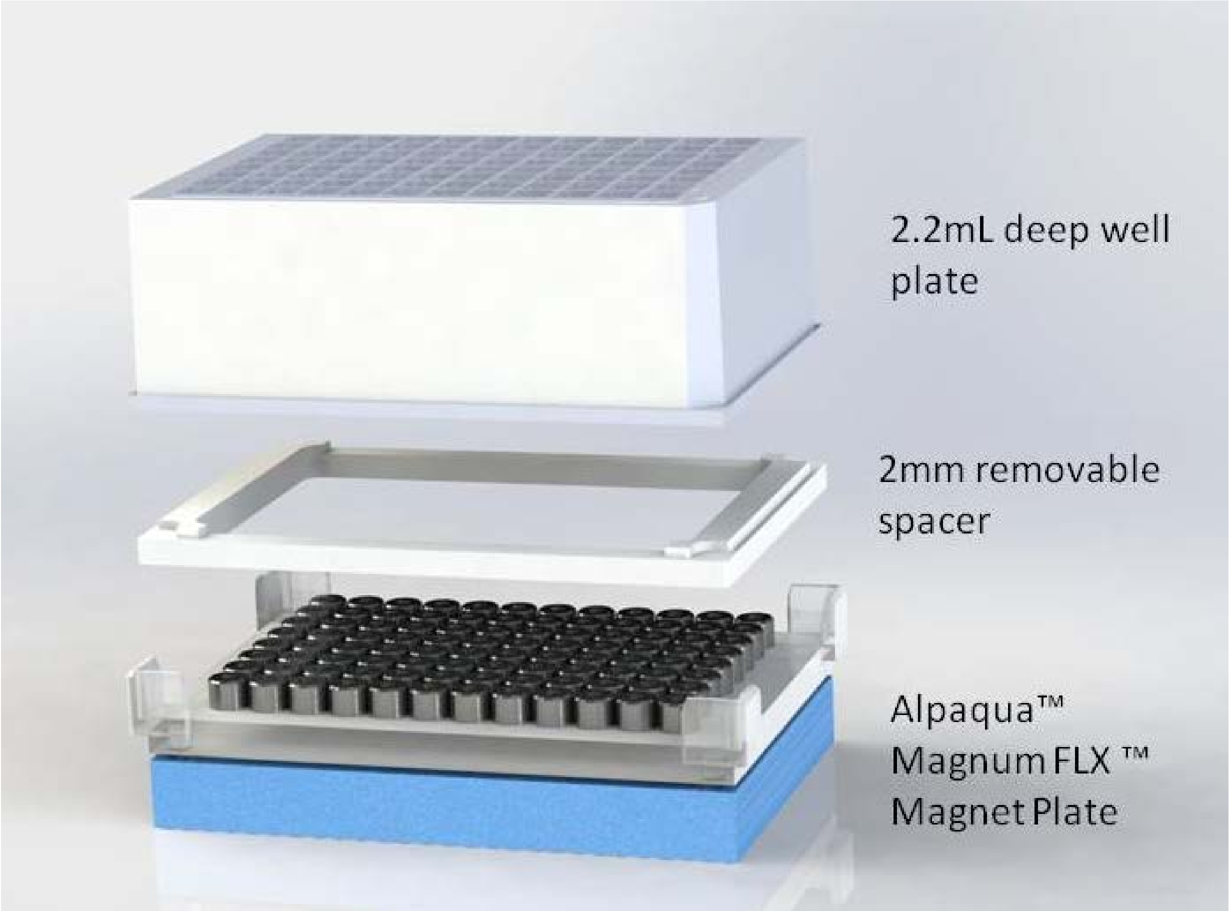


*Use of 2.2 mL plates with a magnet array on the Nimbus required a custom low-profile plate holder for tip clearance. The method runs without an on-deck shaker so we modified bead-elution to accommodate the altered shape and increased surface area of the bottom of the wells. Specifically, the elution volume (40 µL) did not cover the bead ring formed around the inner wall of the well. We 3-D printed a spacer that is inserted on top of the magnet to increase the space between the magnet and plate thus pulling the bead pellet to the bottom of the wells.
