## Appendix 3 to 6 - Figures for "Optimization of magnetic bead-based nucleic acid extraction for SARS-CoV-2 testing using readily available reagents"

A

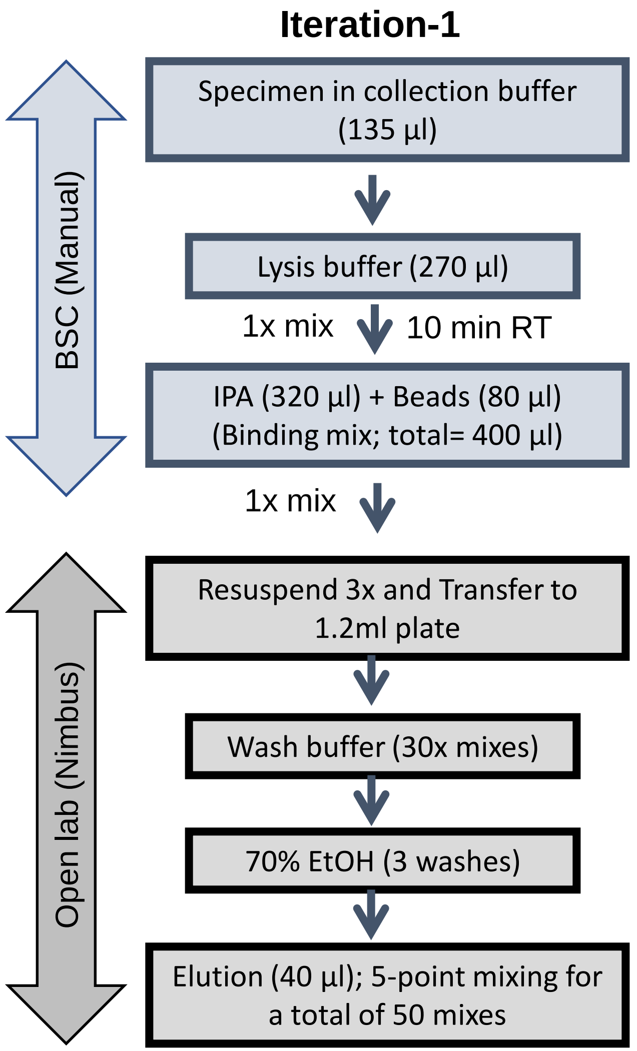


B

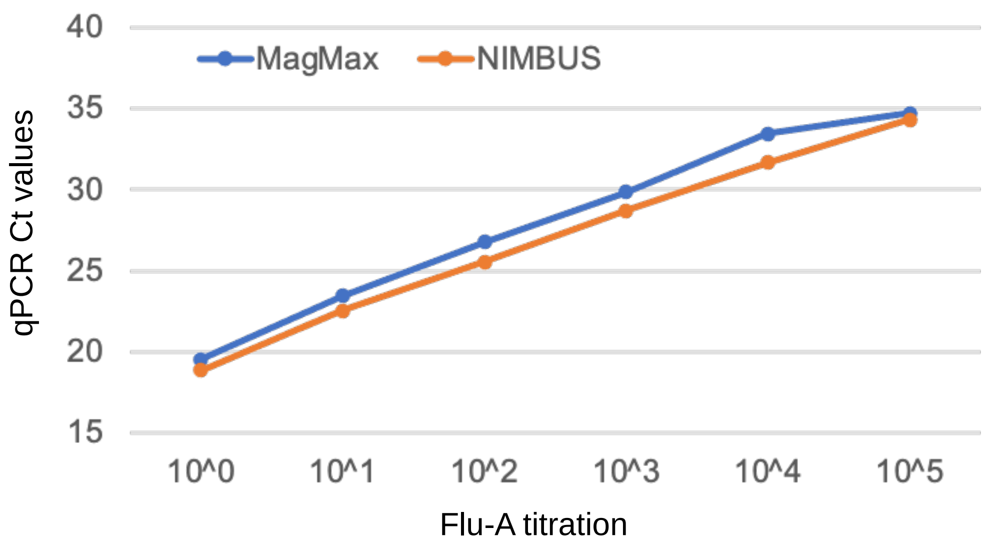


C

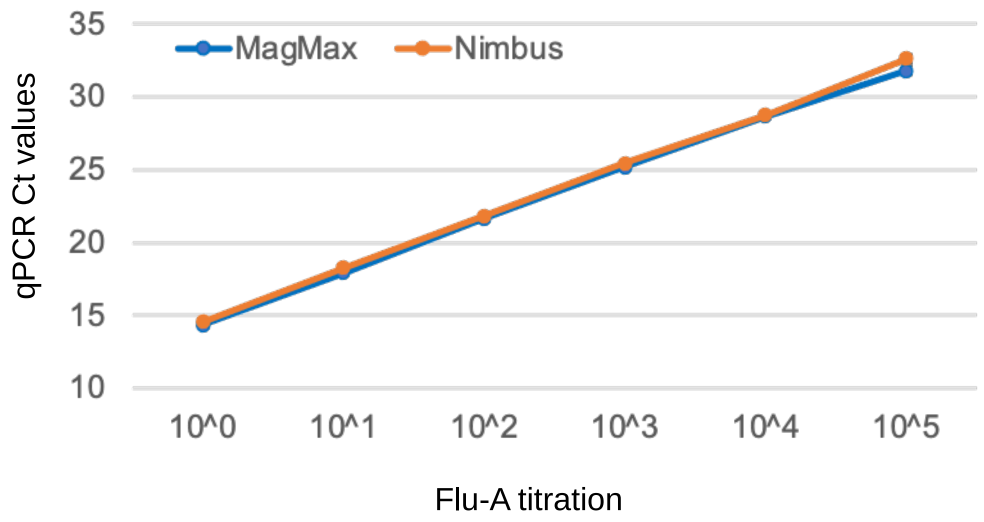


**Appendix 3**

### Iteration-2

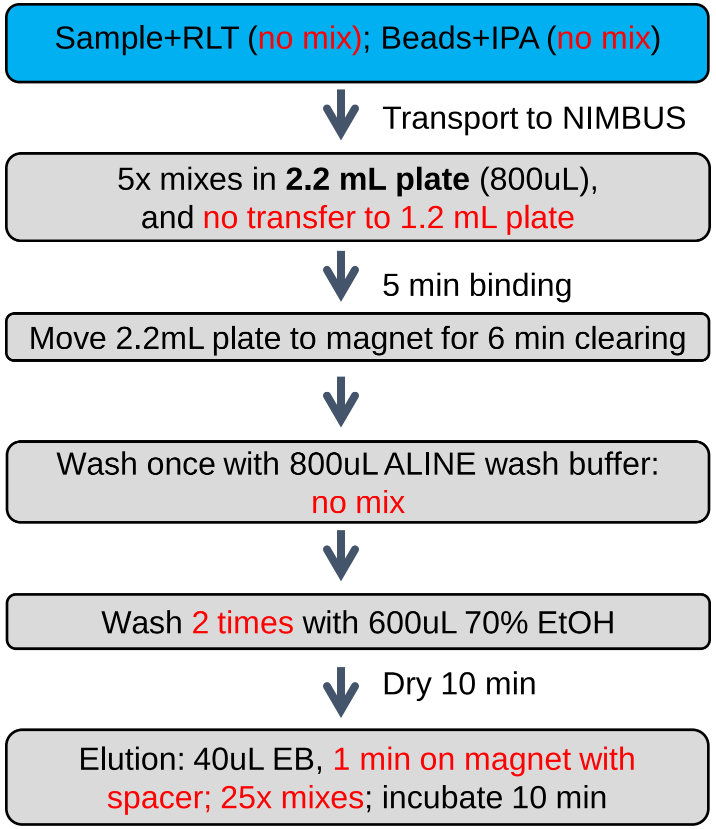


**Appendix 4**

| Step | Sub-Step | Iteration-1 | Iteration-2 | Remark |
|---|---|---|---|---|
| Bead-sample mixture | Liquid Class | DfFS | Jet dispense (JetNoBO) | Jet dispense allows higher dispense speed than aspirate speed, which results in better clearing after mixing |
| | Pause to clear tips after mix | No | Yes (2min) | Allows residual liquid from the surface area for the 1ml tips with 800 µL volume to settle to bottom of the tips |
| | Clearing time on magnet | 6 min | 10 min | |
| | Aspirate supernatant off plate on magnet | Static | Moving aspirate (dllCustAsp) | |
| Aline Wash Buffer | Conditioning | No | Yes | Prevents percolating which results in hanging droplets |
| | Pod stays with in wells when wash buffer is removed | No | Yes | |
| Ethanol Wash | Change tips between washes | No | Yes | |
| | Pod stays with in wells when wash buffer is removed | No | Yes | |
| Elution | Mixing | 5-point X10=50 times | 25 times | |
| | Aspirate to transfer to destination | Static | Moving | |
| | Dispense to destination | Static | Moving | |
| All steps | Slower later pod movement | No | Yes | |
| | Dispense of waste with trailing air gap | No | Waste | Trailing air gap reduces possible hanging waste droplets |
| | Disposal of tips | Vibration | waste Chute | No vibration means less possible aerosols |

**Appendix 5**

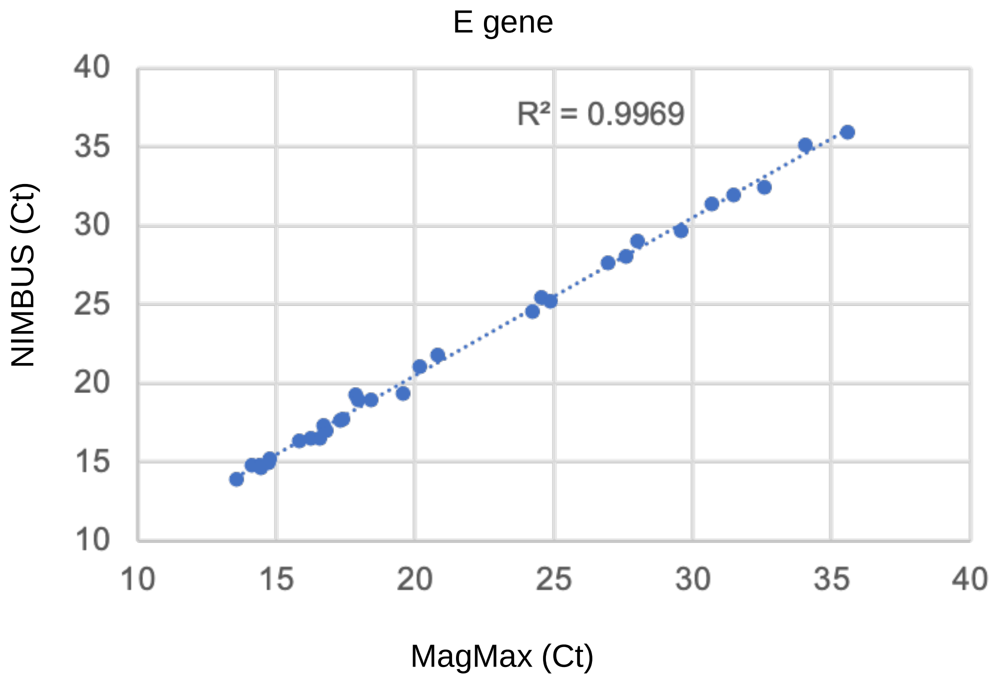
E gene
R² = 0.9969
40
35
30
25
20
15
10
10
15
20
25
30
35
40
NIMBUS (Ct)
MagMax (Ct)

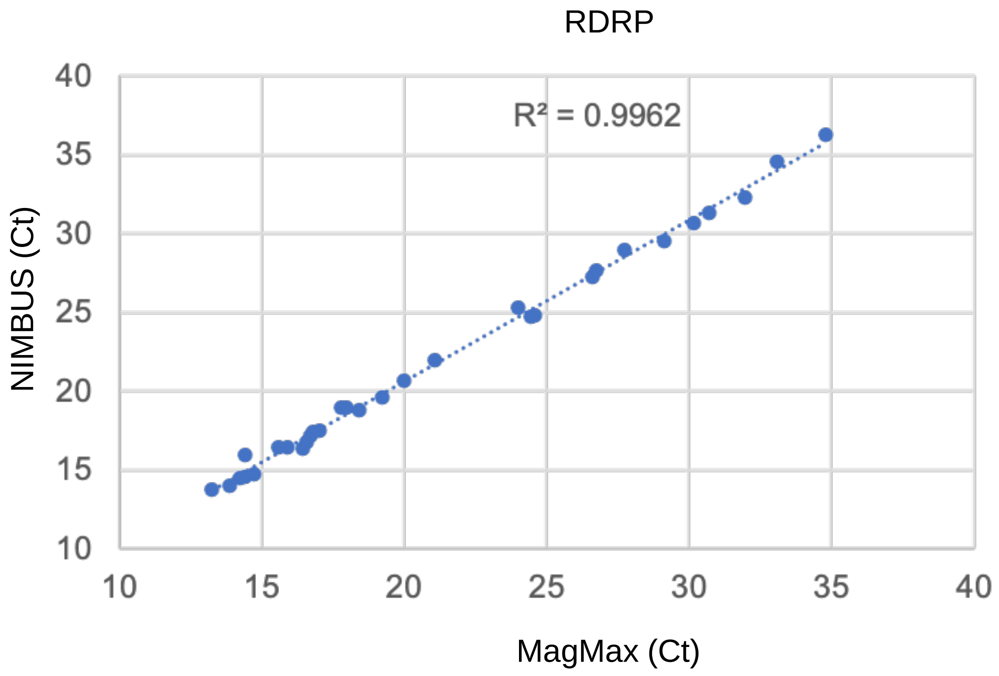
RDRP
R² = 0.9962
40
35
30
25
20
15
10
10
15
20
25
30
35
40
NIMBUS (Ct)
MagMax (Ct)

**Appendix 6**
